# Economic evaluation of participatory women’s groups scaled up by the public health system to improve birth outcomes in Jharkhand, eastern India

**DOI:** 10.1101/2022.09.09.22279761

**Authors:** Hassan Haghparast-Bidgoli, Amit Ojha, Rajkumar Gope, Shibanand Rath, Hemanta Pradhan, Suchitra Rath, Amit Kumar, Vikash Nath, Parabita Basu, Andrew Copas, Tanja A.J. Houweling, Akay Minz, Pradeep Baskey, Manir Ahmed, Vasudha Chakravarthy, Riza Mahanta, Tom Palmer, Jolene Skordis, Nirmala Nair, Prasanta Tripathy, Audrey Prost

## Abstract

An estimated 2.4 million newborn infants died in 2020, 80% of them in sub-Saharan Africa and South Asia. To achieve the Sustainable Development Target for neonatal mortality reduction, countries with high mortality need to implement evidence-based, cost-effective interventions at scale. Our study aimed to estimate the cost, cost-effectiveness, and benefit-cost ratio of a participatory women’s groups intervention scaled up by the public health system in Jharkhand, eastern India. The intervention was evaluated through a pragmatic cluster non-randomised controlled trial in six districts. We estimated the cost of the intervention from a provider perspective, with a 42-month time horizon for 20 districts. We estimated costs using a combination of top-down and bottom-up approaches. All costs were adjusted for inflation, discounted at 3% per year, and converted to 2020 International Dollars (INT$). Incremental cost-effectiveness ratios (ICERs) were estimated using extrapolated effect sizes for the impact of the intervention in 20 districts, in terms of cost per neonatal deaths averted and cost per life year saved. We assessed the impact of uncertainty on results through one-way and probabilistic sensitivity analyses. We also estimated benefit-cost ratio using a benefit transfer approach. Total intervention costs for 20 districts were INT$ 15,017,396. The intervention covered an estimated 1.6 million livebirths across 20 districts, translating to INT$ 9.4 per livebirth covered. ICERs were estimated at INT$ 1,272 per neonatal death averted or INT$ 41 per life year saved. Net benefit estimates ranged from INT$ 1,046 million to INT$ 3,254 million, and benefit-cost ratios from 71 to 218. Our study suggests that participatory women’s groups scaled up by the public health system are highly cost-effective in improving neonatal survival and have a very favourable return on investment. The intervention can be scaled up in similar settings within India and other countries.

## Introduction

There has been a significant decline in the number of neonatal deaths globally, from 5 million in 1990 to 2.4 million in 2020. However, the reduction in deaths during the neonatal period has been slower than that observed for the post-neonatal period [1]. In addition, 80% of neonatal deaths occur in sub-Saharan Africa and South Asia, with neonatal mortality rates (NMR) of 27 and 25 deaths per 1,000 live births, respectively, in 2020 [1]. To achieve the global Sustainable Development Target of neonatal mortality of no more than 12 per 1000 livebirths by 2030 [2], countries with high NMR need to implement evidence-based, cost-effective interventions at scale [3, 4].

Community mobilisation through women’s groups practising Participatory Learning and Action (PLA) is recommended by the World Health Organisation’s (WHO) Global Strategy for Women’s, Children’s and Adolescents’ Health[5] as a cost-effective intervention to improve neonatal survival. This strategy engages communities in a series of monthly meetings following the four phases of PLA: identifying and prioritising health problems in the perinatal period, identifying feasible strategies to address these issues, implementing the strategies, and evaluating the process [6]. Participatory women’s groups have been implemented in small- to medium-scale efficacy trials in Nepal, India, Bangladesh and Malawi[7-11], with several trials demonstrating significant cost-effective reductions in neonatal mortality[12-14].

To accelerate reduction in neonatal mortality in India, where around a fifth of all neonatal deaths happen each year[15], India’s National Health Mission (NHM) in 2016 requested 10 states to consider scaling up participatory women’s groups. This decision was informed by results from two previous efficacy trials in Jharkhand and Odisha [9, 11], a meta-analysis of seven trials of participatory women’s groups[16] and the WHO recommendation [6]. Between 2016 and 2020, with support from Jharkhand NHM, participatory women groups were scaled up across 24 districts of Jharkhand state, covering a population of around 30 million[17, 18]. The programme was called FLAG (Facilitated Learning and Action Groups) and was delivered by public health system frontline government workers called Accredited Social Health Activists (ASHAs). The programme’s effects were evaluated through a pragmatic cluster non-randomised controlled trial in six of Jharkhand’s 24 districts. The trial found a 24% reduction in neonatal mortality, including 26% among the most deprived[19].

This study aimed to assess the cost and cost-effectiveness of the FLAG programme at scale.

## Methods

### Study setting and participants

Jharkhand is a state of Eastern India with a population of around 40 million (estimated for 2021), around 76% of which is rural. Indigenous communities (Scheduled Tribes) constitute 26% of the population[20]. The maternal mortality ratio and neonatal mortality rate in Jharkhand state are 76 per 100,000 livebirths and 33 per 1000 livebirths, respectively[21, 22]. All women living in the programme districts were eligible to take part in participatory women’s groups run under the FLAG programme. The programme’s impact evaluation included women of reproductive age (15-49 years) who gave birth during the evaluation period (i.e. 1^st^ September 2017 - 31^st^ August 2019) in six purposefully selected districts.

### FLAG Intervention

The FLAG programme was implemented by the National Health Mission (NHM), Jharkhand, in partnership with Ekjut, a civil society organisation working with women’s groups in Jharkhand since 2006. The programme began in six blocks of six districts in 2015-6, and gradually expanded to 21 of Jharkhand’s 24 districts from 2017 onwards. The three remaining districts did not receive the intervention until 2019 and were ‘comparison’ districts for the programme’s impact evaluation.

A detailed description of the women’s group intervention is reported elsewhere [19]. In brief, the intervention comprised monthly women’s group meetings (around 36 monthly meetings) following a Participatory Learning and Action (PLA) cycle. Meetings were facilitated by ASHAs and their supervisors, who supervised approximately 15-20 ASHAs. The women’s groups intervention followed three phases of a PLA cycle. In first phase, group members identified and prioritised maternal and newborn health problems in their community. In phase two, they discussed the causes of problems prioritised in the first phase and identified locally feasible strategies to address the causes. During this phase, groups organised a larger community meeting to share their prioritised health problems and the strategies they had identified with the wider community. In the third phase, groups implemented their strategies. In the fourth phase, groups evaluated the overall cycle. In each phase, different participatory techniques such as voting to prioritise problems, storytelling and games were used to facilitate discussion.

Women’s groups mainly focused on health problems in the perinatal period, but a number of extra meetings were added to phase three, focusing on issues such as infant and young child feeding, maternal nutrition and family planning. ASHAs, in addition to leading monthly PLA meetings, were also required to carry out their routine work in the community, including promoting antenatal care and institutional delivery, and providing postnatal home visits as part of the Home-Based Newborn Care programme [23, 24].

With support from Ekjut, NHM District and Block-level staff gave ASHA Facilitators three rounds of training on PLA, with each training lasting five days. ASHA Facilitators then provided on-the-job training to all ASHAs in their catchment area (around 20 villages and 20 ASHAs), using an odd-even approach covering 10 villages each month. We described this low-cost training method in detail elsewhere[19]. ASHA Facilitators received an incentive of INR 1,000 (USD 13) to conduct 10 meetings per month, and ASHAs received INR 100 (USD 1.3) per meeting. ASHAs and ASHA Facilitators formed around one group per 1000 population and encouraged pregnant women and women from vulnerable groups such Scheduled Tribes, Scheduled Castes and remote hamlets, to participate in group meetings. ASHA Facilitators and ASHAs also attended monthly review meetings with district-level coordinators as well as biannual meetings at the state-level to supplement their training.

### Impact evaluation

We used a pragmatic cluster non-randomised controlled trial design to evaluate the effects of FLAG. Three of the six evaluation districts (Ranchi, West Singhbhum, Khunti) started women’s groups in 2017, while the other three (Bokaro, Palamu, Dumka) started in 2019. The allocation of districts to a 2017 or 2019 start was purposive. The evaluation team collected data on births, deaths and perinatal practices among women of reproductive age prospectively in 100 purposively selected units of 10,000 population, each located in six purposefully selected districts. We collected data during a six-month baseline period (1^st^ March-31^st^ August 2017) and a 24-month evaluation period (1^st^ September 2017-31^st^ August 2019). The trial’s primary outcome was neonatal mortality. Secondary outcomes included stillbirths, perinatal mortality, maternal mortality, as well as preventive and care-seeking practices for women during the perinatal period [19]. We used an intention-to-treat approach and included data from all eligible mothers in the evaluation area. All trial analyses were carried out a cluster-level, with districts as clusters. All models were adjusted for tribe/caste, maternal schooling, maternal literacy, asset quintile, and baseline values for each outcome. More details on the evaluation design and analysis are provided elsewhere[19]. The trial found a 24% reduction in neonatal mortality (Adjusted Odds Ratio [AOR]: 0.76, 95% CI: 0.59-0.98) in districts that began the intervention in 2017 (‘early districts’) compared to those than began in 2019 (‘delayed districts’)[19].

### Impact at scale and life years saved

We extrapolated the effect of the intervention on neonatal mortality found in the six trial districts to other districts in the State that met three conditions: received all three PLA trainings; completed two-thirds or more of perinatal meetings by 2019 and; had 30% or more of pregnant women in ASHA catchment areas participating in group meetings. Twenty out of 24 districts met these criteria. We estimated the number of neonatal lives saved in rural areas of 20 districts using the following formula:

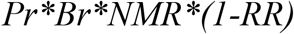

where Pr is rural population in 20 districts, Br is birth rate in crude birth rate in rural area, NMR is neonatal mortality rate in rural areas without intervention, and RR is the Risk Ratio for the intervention in the evaluation districts[19]. We then estimated life years saved by multiplying the number of neonatal deaths averted in 20 districts by 30.81, which corresponds to a standard life expectancy of 86 years, as recommended by Global Burden of Diseases 2010 [25], discounted at 3%.

### Intervention costing

We estimated the cost of the intervention implemented in 20 districts from a programme provider perspective. The costing’s time horizon was 42 months (October 2016-March 2020). A combination of activity-based, expenditure and ingredient approaches[26] were used to estimate the costs. We collated financial costs data retrospectively from the expenditure reports and project accounts of implementing agencies, namely Ekjut and NHM Jharkhand. In addition, we used project records on the number of meetings held by ASHAs and incentive data (i.e. 100 INR per meeting) to estimate the total incentives paid to ASHAs. Cost data were input into an excel-based tool and categorised according to line item, intervention activities, and implementing agency (Ekjut and NHM). Table 1 provides a description of cost categories. Staff costs were allocated to different activities using staff time-use data collected through consultation with the project staff. We excluded all research costs as these were primarily incurred by the impact evaluation.

**Table 1:** FLAG Intervention costs by input and activity/intervention component: definitions^1^

| Description |  |
| --- | --- |
| **Costs by line item** |  |
| Staff | Value of staff time contributed to the intervention |
| Materials | Costs of materials related to running PLA meetings or trainings including costs of ASHA Module and picture cards printing |
| Travel | Travel costs, including per diem and other allowances |
| Overhead/joint | Include field offices' running costs and rent, utilities, communications and recruitment costs |
| Mixed inputs | Training sessions and meetings, where different inputs are utilised. |
| Other | Professional fees for Video Documentation of training sessions |
| **Costs by activity/component** |  |
| Trainings–PLA | Training to ASHAs and ASHA supervisors |
| Trainings–other | Project staff capacity building and training workshops for different government agencies |
| PLA implementation | Incentives to ASHAs (n=39,000) and ASHA facilitators/supervisors (n=1,877) and other costs of running PLA groups including picture card and module printing |
| Monitoring & Evaluation | Includes costs of setting up monitoring information system/dashboard, monthly review meetings and data entry Cost |
| Admin/Joint | Overhead cost and time spent in administration/joint activities by staff |
| Preparation/Planning | Include project planning meetings, district level orientation and community sensitisation meetings, and recruitment |
| Coordination | State level progress meetings and liaising with state health offices. It includes costs of staff who allocated a substantial proportion of their times to coordination activities: State programme coordinator<sup>1</sup> (n=1), District Programme Coordinators<sup>2</sup> (n=21), District Coordinators (n=18)<sup>3</sup>, Government Partnership Officer (n=1)<sup>3</sup>, Regional Coordinators (n=5)<sup>3</sup>, Programme Officer (n=1)<sup>3</sup>, Communication officer (n= 1)<sup>3</sup>, Scale up Manger (n=1)<sup>3</sup>, Programme Coordinator (n=1)<sup>3</sup>. |

<sup>1</sup>PLA: Participatory Learning and Action; ASHAs: Accredited Social Health Activists. <sup>2</sup>Funded by National Health Mission, Jharkhand <sup>3</sup>Funded by Ekjut

### Analyses

All costs were adjusted for inflation, discounted at 3% per year as recommended by WHO-CHOICE[27] and the Gates/iDSI Reference Case for Economic Evaluation[28], then converted to 2020 international dollars (INT$) using the Purchasing Power Parity conversion factor of 21.2 for India[29]. Incremental cost effectiveness ratios (ICERs) were estimated using the outcome data from the effect at scale analysis and presented in terms of cost per neonatal death averted and cost per life year saved. We have followed recommendations from the Gates/iDSI Reference Case for Economic Evaluation[28] and Consolidated Health Economic Evaluation Reporting Standards (CHEERS) statemen [30] in our analysis and reporting.

In addition to the ICER, we also estimated unit costs for the intervention in terms of total and average annual costs per livebirth covered by the intervention.

#### Sensitivity analysis

We conducted a series of deterministic one-way sensitivity analyses and probabilistic sensitivity analyses (i.e. a Monte Carlo simulations). We quantified the impact of the following parameters in one-way sensitivity analyses: intervention effectiveness (95% confidence interval), intervention cost (+/- 25%), discount rate (differential rate for cost and outcome −0%-10%), life expectancy (standard 86 years vs local life expectancy and WHO Global Health Estimates), baseline neonatal mortality rate (NMR), and rural crude birth rate. In the probabilistic sensitivity analyses, we tested simultaneous variations of multiple parameters, using their assumed distribution, generating a total of 1000 iterations in Excel. Table S1 in the annex shows all the parameters that were varied in sensitivity analyses, alongside results from probabilistic sensitivity analysis.

#### Benefit cost analysis/ Return on investment

We estimated the value of avoided neonatal deaths following the benefit transfer approach suggested by Robinson et al[31]. Robinson et al[31] recommended following three alternative approaches: (1) extrapolating a value of statistical life (VSL) from the US (US$ 9.4 million in 2016, equivalent to 160 times Gross National Income-GNI per capita) with an income elasticity of 1.5; (2) estimating VSL as 160 times GNI per capita, with an income elasticity of 1.0 (US base value); (3) estimating VSL as 100 times GNI per capita, with an income elasticity of 1.0 (OECD base value). We used GNI per capita of 2015 INT$ 6060 (or US$ 1600) for India in our calculations.

We also estimated a constant value of statistical life year (VSLY) as recommended by Robinson et al[31]. VSLY is estimated by dividing the VSL by the undiscounted life expectancy of an adult of average age. We used the life expectancy of a 40-44 years old adult in India in 2019 (35.4 years), based on the life table for India in 2019 [32]. We then multiplied the VSLY by the total neonatal life years saved to estimate total mortality benefits.

VSL reflects an individual’s willingness to pay for a change in his/her own risk of mortality (or life expectancy in case of VSLY) within a specific time period (usually a year)[31]. Income elasticity represents association between VSL and income. An income elasticity of greater than one suggests that the ratio of VSL to GNI per capita is smaller in lower income compared to higher-income populations [31].

To be conservative, we used the VSLY calculated from approach (1), i.e. 160 times GNI per capita with an income elasticity of 1.5, with benefits discounted at 10%, as our base case scenario. We used discount rates of 3%, 5% and 10% (base-case) for benefit estimates as recommended by iDSI reference case [28] and Haacker et al [33]. We estimated net benefits by subtracting total intervention costs from total monetised mortality benefits, and the benefit-cost ratio (BCR) by dividing total monetised benefits by total intervention cost.

#### Ethics

Ethical approval for the FLAG programme evaluation was obtained from an independent ethics research committee linked to Ekjut in Ranchi, and University College London’s Research Ethics Committee (Reference: 1881/003). We sought individual informed consent from all participants in the evaluation study.

## Results

The total and annual cost of programme implementation in 20 districts were INT$ 15,017,396 and INT$ 4,433,165, respectively. Intervention covered around 1.6 million livebirths across 20 districts which translated to INT$ 9.4 per livebirths (Table 2).

**Table 2:**
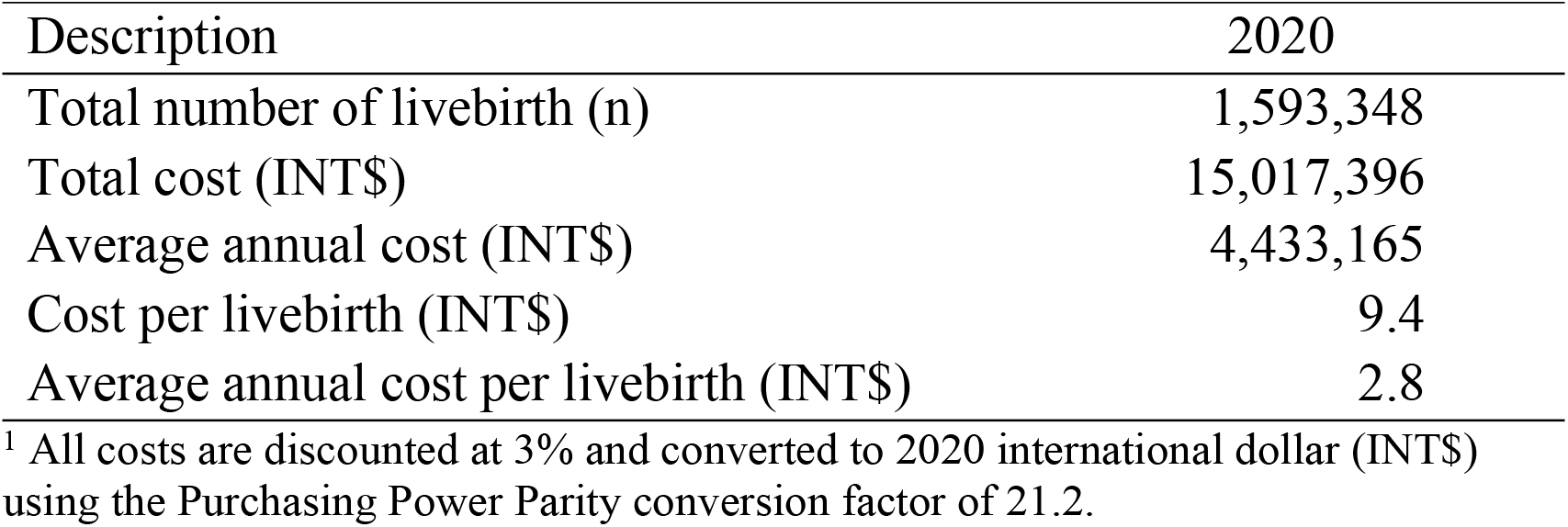
Total, annual and unit costs of the FLAG intervention^1^

Staff costs constituted around 80% of all intervention costs. Around 40% of staff costs were incentives for ASHAs and ASHA facilitators. Mixed inputs (mainly trainings and meetings) were the second largest cost category, amounting to 13% of all costs (Table 3 and Figure 1). PLA implementation (which mainly include incentives paid to ASHAs and ASHA facilitators), and training constituted around 80% of all intervention costs. Preparation, planning and community sensitisation meetings and recruitment constituted less than 1% of total costs (Table 3 and Figures 2).

**Table 3:**
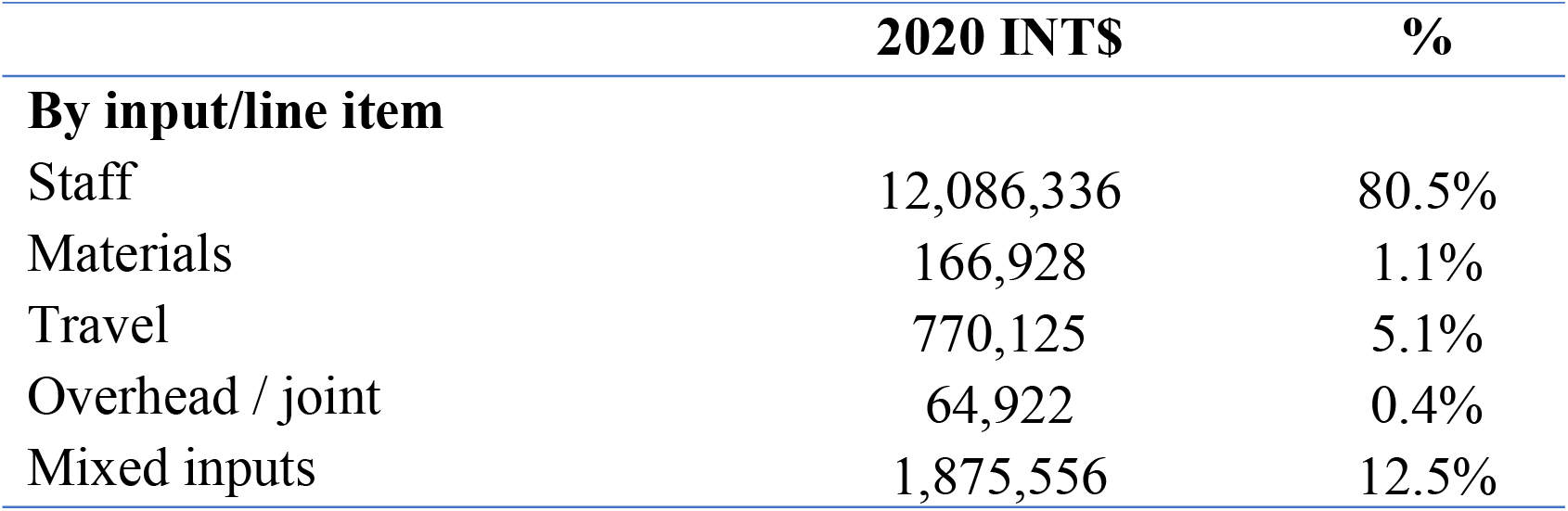
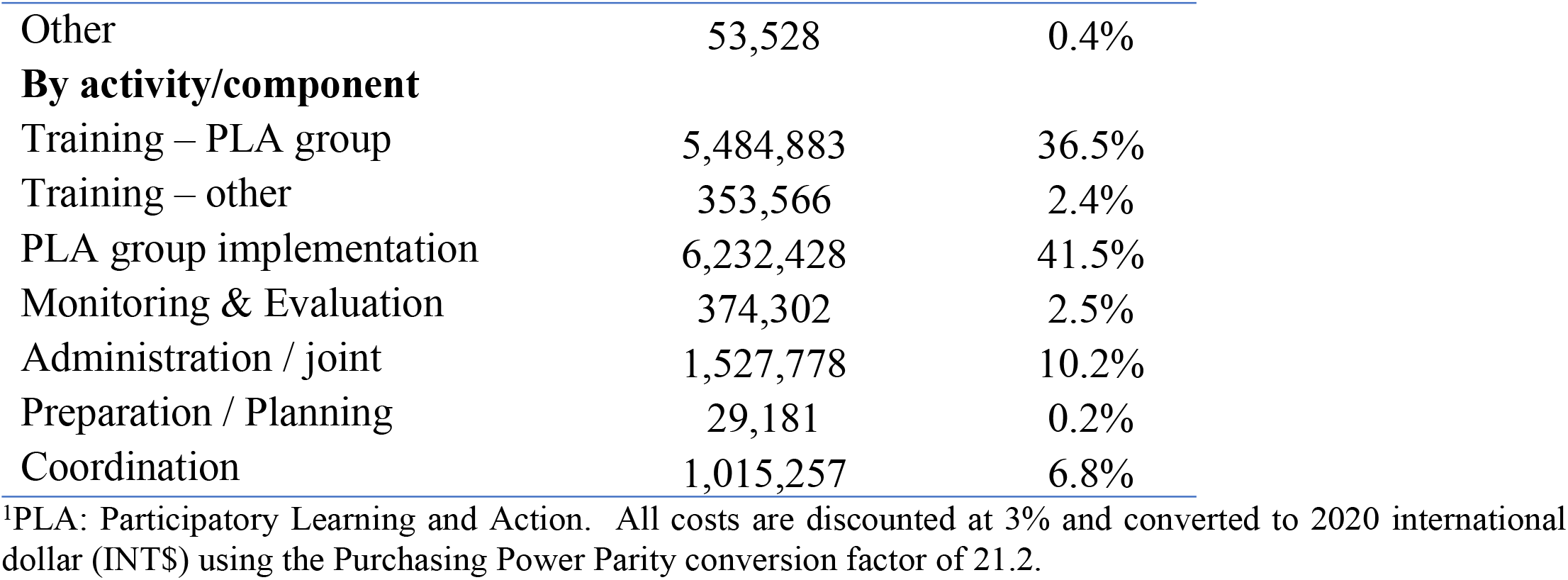
Total FLAG intervention costs by cost category^1^

**Figure 1:**
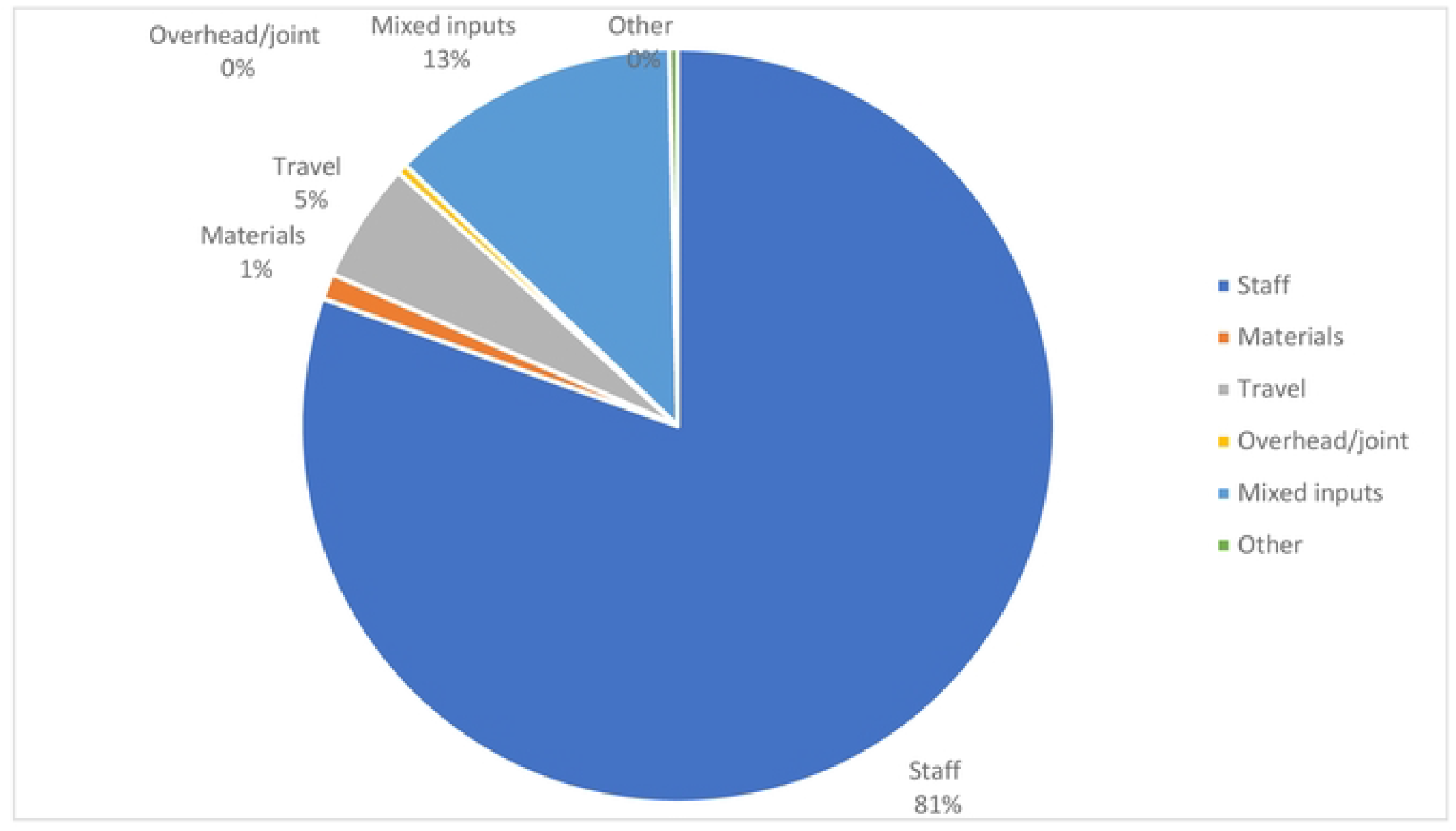
FLAG intervention costs, by input

**Figure 2:**
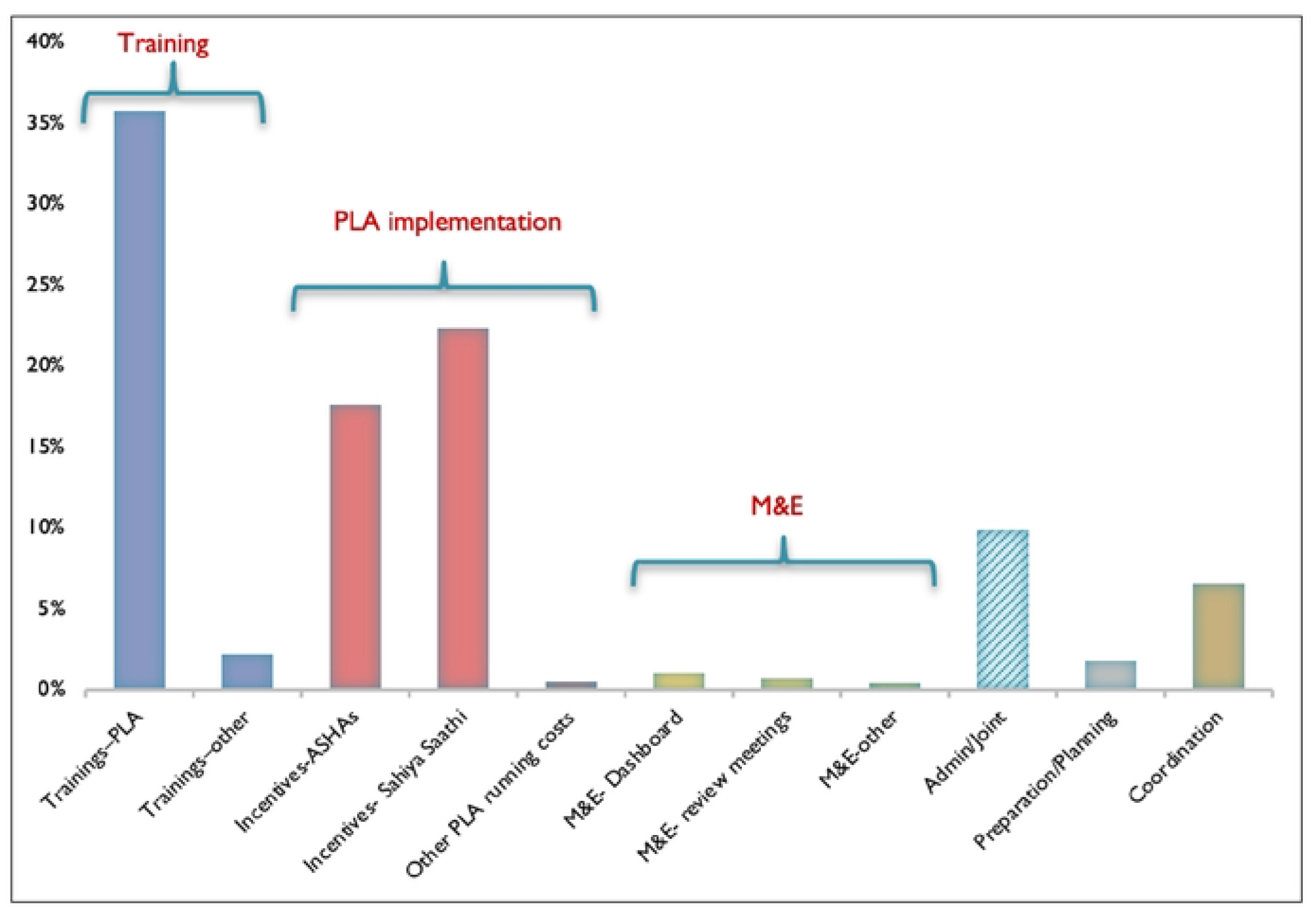
FLAG intervention costs, by activity/component

FLAG’s impact evaluation found that the intervention saved 188 neonatal lives over a two years period (min: 16 – max: 327), using lower and higher bound of confidence interval around impact) in the three early intervention districts taking part in the evaluation[19]. Over the 42 months of scale up, we estimated that the intervention saved a total 11,803 neonatal lives across 20 districts (min: 1026 - max: 20,527) [19]. This translates to an estimated 363,621 (discounted) life years saved, with a range of 31,609 to 632,386 (Table 4).

**Table 4:** Effectiveness and cost-effectiveness of the FLAG programme

| **Description** | **Base-case** | **Lower impact** | **Higher impact** |
| --- | --- | --- | --- |
| Neonatal deaths averted (N) | 11,803 | 1,026 | 20,527 |
| Life years saved, undiscounted (N)<sup>1</sup> | 1,015,036 | 88,264 | 1,765,280 |
| Life years saved, 3% discounted (N)<sup>2</sup> | 363,621 | 31,609 | 632,386 |
| Cost per neonatal death averted (2020 INT\$)<sup>3</sup> | \$1,272 | \$14,632 | \$732 |
| Cost per neonatal life-year saved (2020 INT\$) | \$41 | \$475 | \$24 |
| GDP per capita, India, (2019INT\$) | \$7,034 | | |
| ICER as % GDP per capita | 0.6% | 6.9% | 0.3% |

<sup>1</sup> Using standard life expectancy of 86, <sup>2</sup> multiplied by 30.81, which corresponds to a standard life expectancy of 86 years, discounted at 3%. <sup>3</sup>All costs are discounted at 3% and converted to 2020 international dollar (INT\$) using the Purchasing Power Parity conversion factor of 21.2.

ICERs were estimated as INT$ 1,272 (range: INT$ 732-INT$ 14,632) per neonatal death averted or INT$ 41 (range: INT$ 24-INT$ 475) per life year saved. Costs per life year saved ranged from 0.3% to 7% of GDP per capita of India (INT$ 7,034 in 2019), indicating that the intervention was highly cost-effective when compared against the common GDP-based cost-effectiveness thresholds such as Woods et al[34] and WHO CHOICE[35].

### Sensitivity analysis

Figure 3 presents results from one-way deterministic sensitivity analyses. The results indicated that uncertainties around intervention effectiveness, NMR in areas without intervention, differential discount rates and rural crude birth rates led to significant variation in the ICER. For example, the uncertainty around intervention effectiveness (95% confidence interval risk ratio: 0.60-0.98), led to variations of −43% to 1050% in ICER, or INT$24 to INT$475 (Table S1). Varying NMR rate in areas without intervention (from 18 to 52 per 1000 live births) led to variations of −38% to +76% in the ICER (i.e. INT$ 25 - INT$73). In contrast, using local life expectancy of 70 or 91.9 used by WHO Global Health Estimates had only small effects on the cost effectiveness results (+5% and −1%, respectively). The 95% Confidence Interval around the ICER calculated from 1000 iterations in PSA was INT$32 to INT$ 62 (Figure S1). Overall, varying uncertain parameters did not change the conclusion that the intervention is highly cost-effective.

**Figure 3:**
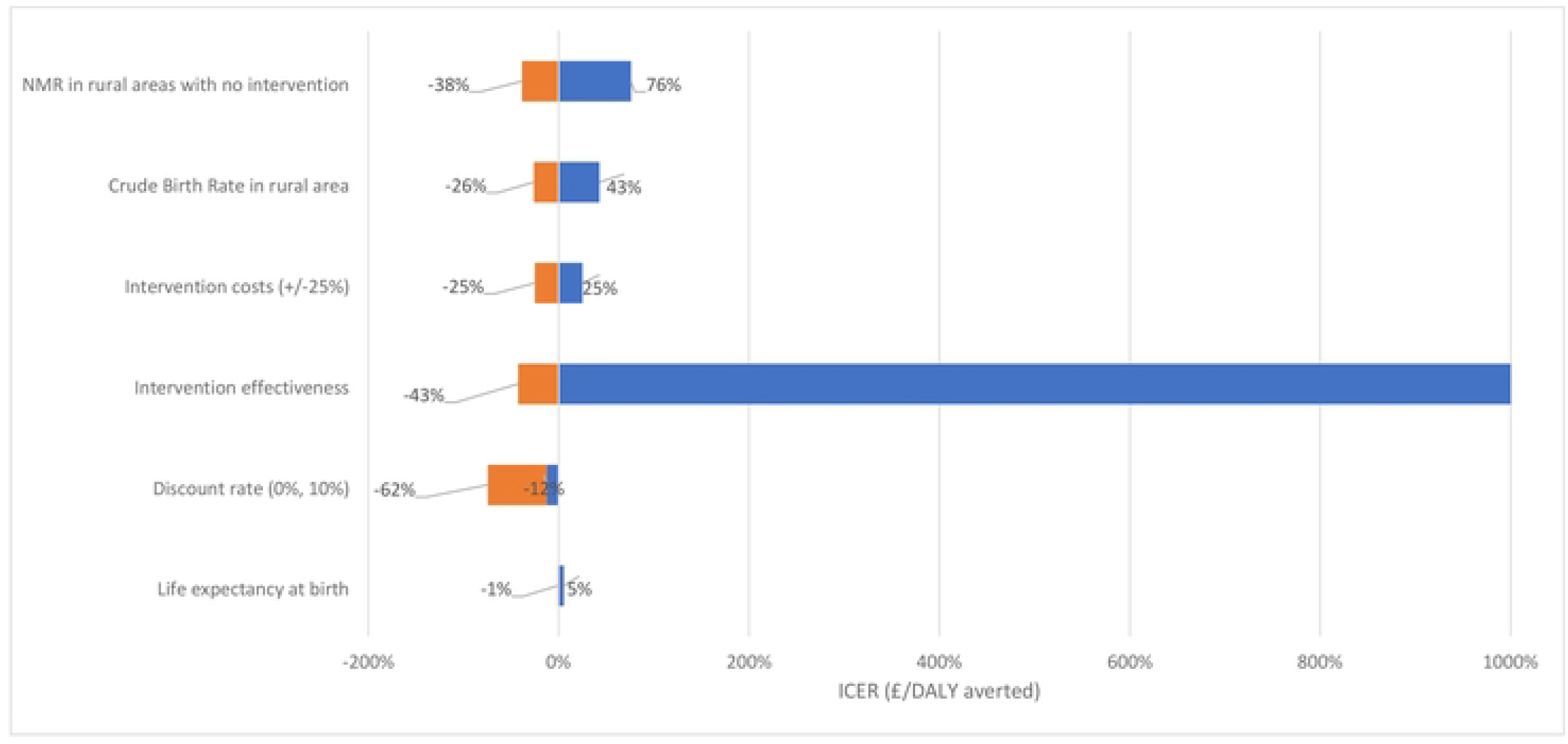
Tornado diagram of percentage change in the incremental cost-effectiveness ratio (ICER) resulted from deterministic one-way sensitivity analysis of key parameters

#### Benefit cost analysis

Table 5 presents the results of the VSL, VSLY estimates. Table 6 shows total mortality benefits estimations. Estimated total benefits due to aversion of premature mortality, using US-VSLY extrapolation approach, ranged from INT$ 1,061 million with a 10% discount rate to INT$ 3,269 million with a 3% discount rate. The net benefit estimates ranged from INT$ 1,046 million (base case) to INT$ 3,254 million, and BCRs from 71 (base case) to 218.

**Table 5:** Estimates for VSL and VSLY for India (2015 INT$)^1^

|  | **Approach 1: GNI**<br>**per capita X 160**<br>**(elasticity 1.5)** | **Approach 2: GNI**<br>**per capita X 160**<br>**(elasticity 1)** | **Approach 3: GNI**<br>**per capita X 100**<br>**(elasticity 1)** |
| --- | --- | --- | --- |
| **VSL** | 318,287 | 969,600 | 606,000 |
| **VSLY** | 8,991 | 27,390 | 17,119 |

<sup>1</sup>VSL: value of a statistical life; VSLY: value of statistical life year; GNI: Gross National Income; INT\$: International Dollar

**Table 6:** Total mortality benefits estimates for FLAG, using approach 1^1^

|  | **3% discounted** | **5% discounted** | **10% discounted** |
| --- | --- | --- | --- |
| **VSL<sup>2</sup>**(lower-higher impacts)-<br>2015 INT\$, million | 3,756 (326–6,533) | 3,756 (326–6,533) | 3,756 (326–6,533) |
| **VSLY<sup>3</sup>**(lower-higher impacts)-<br>2015 INT\$, million | 3,269 (284–5,687) | 2094 (182–3,641) | 1,061 (92–1,846) |
| **Net Benefit** |  |  |  |
| VSL (2015 INT\$, million) | 3,742 (311–6,518) | 3,742 (311–6,518) | 3,742 (311–6,518) |
| VSLY (2015 INT\$, million) | 3,254 (269–5,671) | 2,079 (167–3,626) | 1,046 (77–1,831) <sup>4</sup> |
| **BCR** |  |  |  |
| VSL (lower-higher impacts) | 250 (22–435) | 250 (22–435) | 250 (22–435) |
| VSLY (lower-higher impacts) | 218 (19–379) | 139 (12–242) | 71 (6–123)*** |

<sup>1</sup>VSL: value of a statistical life; VSLY: value of statistical life year; GNI: Gross National Income; BCR: benefit-cost ratio; INT\$: International Dollar. <sup>2</sup>Estimated as total death averted multiply by VSL of INT\$ 318,287. <sup>3</sup> Estimated as total life years saved multiply by VSLY of INT\$ 8,991. <sup>4</sup>Base-case scenario.

## Discussion

Our study is the first to assess the cost and cost-effectiveness of a large-scale community mobilisation intervention through participatory women’s groups, delivered by a public health system, to improve neonatal survival. Our findings suggest that the scaled-up intervention was highly cost-effective in reducing neonatal mortality and has a very favourable return on investment. The incremental cost-effectiveness ratio of $INT41 is substantially lower than India’s GDP per capita (ranging from 0.3% to 7% of GDP). FLAG’s ICER is 85% lower than that found in a smaller-scale efficacy trial of women’s groups facilitated by ASHAs conducted in Jharkhand and Odisha (ICER of 2017 INT$ 274 or US$83), and 70% lower than in another smaller trial of women’s groups supported by salaried facilitators in the same states (ICER of 2016 INT$ 135) [13] [12]. Similarly, FLAG’s ICER is also substantially lower than those reported for small scale efficacy trials of women’s groups in Nepal, Bangladesh, and Malawi[12] (Table S2).

A similar conclusion can be drawn by comparing FLAG results with two small (JEEViKA-MC) [36] and large (Parivartan) scale[37] implementation studies of Self-help groups (SHGs) with maternal and child health components. The Parivartan programme [37] (2013 - 2016) was implemented in eight districts of Bihar (55 blocks) and reached an estimated 275,000 women of reproductive age. The programme aimed to improve maternal and newborn health and sanitation behaviours. It cost 2016 US$11 per woman reached and US$3,825 per life year saved, ranging from US$3,221 to US$11,731 per life year saved, as compared to the

INT$ 41 per life year saved in FLAG. The JEEViKA Multisectoral Convergence Model (JEEViKA-MC) was a pilot project implemented between 2016 and 2018 (27 months) in 12 Gram Panchayats (villages) of Saharsa, Bihar to address the underlying causes of undernutrition among women and children. The project, which was developed by the Bihar Rural Livelihoods Promotion Society with support from the World Bank, builds on the JEEViKA model, a livelihood programme that uses SHG as a platform and aims to integrate health and nutrition education into existing SHG meetings. The target population were SHGs members, with a particular focus on households with young children, mothers of young children, and pregnant women. The intervention covered 1,591 SHGs and 3,823 target beneficiaries (women and children) at a cost of 2018 US$62 per beneficiary[36]. Although direct comparisons are impeded by different definitions of beneficiaries and reporting for different years, data suggest FLAG was more cost-effective than these two past programmes.

A combination of factors might have contributed to the reduced costs of FLAG. Employing incentivised ASHA and ASHA facilitators rather than salaried facilitators and supervisors is potentially the main driver of cost savings. The ‘odd-even’ on-the-job-training approach used at scale is another potential driver. The intervention was implemented in a less intensive manner than in previous small scale efficacy in Jharkhand[9] [13]. In FLAG, there was one women’s group per c. 1000 population compared to one per c.500 population in previous trials in India. Finally, the results also potentially reflect economies of scale (See Figure S1); the intervention area covered 1.6 million livebirths at a very low unit cost (cost per livebirths) (INT$ 9.4 compared with average unit cost of INT$ 203, range: 2016 INT$ 61-537)[12] in smaller scale trials of women’s groups (Table S2). As Figure S2 illustrates, unit costs decreased with the increased scale of the intervention, which is in line with conclusion from scale up of SHGs in India, in general [38].

The key drivers of cost-effectiveness, based on our sensitivity analysis, were intervention effectiveness, NMR in areas with no intervention, as well as the discount rate and rural crude birth rate. Sensitivity analyses showed that the intervention is still highly cost-effective in settings with NMR less than 32 deaths per 1000 livebirth, if all other parameters remain constant. This conclusion still stands if a lower NMR and the lower bound of the confidence interval for effectiveness is used (with ICER of INT$ 836). However, groups have not been implemented in rural settings with NMR of less than 20 deaths per 1000 livebirth in India or elsewhere[16], therefore the results for these settings should be interpreted with caution. Our cost-effectiveness results could be transferable to other districts in India or rural settings of other countries with high NMR and similar health system characteristics such as the accessibility of health services.

The BCR of INT$71 to INT$218 (US$62-US$191) for FLAG indicates a large return on investment. This result is in line with evidence from other maternal, newborn and child health (MNCH) interventions[39]. The BCR for FLAG is at the higher end of available BCRs reported for MNCH, alongside childhood nutrition interventions[39]. However, it should be considered that these studies are not directly comparable due to intervention type, methodology used and context of the studies, which mainly are from Sub-Saharan Africa.

Our study has two main limitations. First, we estimated costs from a program provider perspective. We therefore did not include indirect costs/opportunity costs to intervention participants, ASHAs and other community health providers affected by the intervention. Although it would be ideal to estimate full implementation costs from a societal perspective, it was not possible to collect more data due to the retrospective nature of the costing exercise. However, the program provider perspective is reflective of actual costs and budget impact of scaling up the intervention if scaled elsewhere in rural, underserved areas of India or other settings. In addition, including these costs would not change the conclusion that the intervention is highly cost-effective. Second, using neonatal mortality as a primary outcome in our analysis likely underestimates the impact of women’s groups. We have not included potential short and long-term health benefits beyond neonatal period and for mothers, children and their siblings. Observed improvements in exclusive breastfeeding, and postnatal visits might result in long-term health benefits for both mothers and children.

## Conclusion

Our study indicates that participatory women’s groups scaled up by the public health system in India were highly cost-effective in improving neonatal survival. The intervention can be scaled in similar settings within India and other countries.

## Data Availability

Aggregated cost data are provided in the tables within the paper. Trial outcome data are reported in detailed elsewhere (Nair N et al, 2021).

## Acknowledgements

We thank all women participated in the group meetings, and mothers and their family members who participated in impact evaluation interviews. We also thank the ASHA Facilitators and ASHAs of Jharkhand, who delivered the intervention.

## References

1. UNICEF. Levels & Trends in Child Mortality: Report 2020 2020 [cited 2021 15 December]. Available from: https://data.unicef.org/resources/levels-and-trends-in-child-mortality-2020/.

2. McArthur JW, Rasmussen K, Yamey G. How many lives are at stake? Assessing 2030 sustainable development goal trajectories for maternal and child health. BMJ. 2018;360:k373. Epub 2018/02/17. doi: 10.1136/bmj.k373. PubMed PMID: 29449222; PubMed Central PMCID: PMCPMC5813301 interests and have no relevant interests to declare. Provenance and peer review: Not commissioned; externally peer reviewed.

3. Bhutta ZA, Das JK, Bahl R, Lawn JE, Salam RA, Paul VK, et al. Can available interventions end preventable deaths in mothers, newborn babies, and stillbirths, and at what cost? The Lancet. 2014;384(9940):347–70. doi: https://doi.org/10.1016/S0140-6736(14)60792-3.

4. Hug L, Sharrow D, Zhong K, You D, on behalf of the United Nations Inter-agency Group for Child Mortality Estimation. Levels and Trends in Child Mortality: Report 2018. New York: United Nations Children’s Fund, 2018.

5. World Health Organization. Global Strategy for Women’s, Children’s and Adolescents’ Health (2016-2030): World Health Organization; 2010 [cited 2021 15 December]. Available from: https://www.who.int/data/maternal-newborn-child-adolescent-ageing/global-strategy-data.

6. World Health Organization, editor. WHO recommendation on community mobilization through facilitated participatory learning and action cycles with women’s groups for maternal and newborn health. Geneva: World Health Organization; 2014.

7. Manandhar DS, Osrin D, Shrestha BP, Mesko N, Morrison J, Tumbahangphe KM, et al. Effect of a participatory intervention with women’s groups on birth outcomes in Nepal: cluster-randomised controlled trial. The Lancet. 2004;364(9438):970–9.

8. Azad K, Barnett S, Banerjee B, Shaha S, Khan K, Rego AR, et al. Effect of scaling up women’s groups on birth outcomes in three rural districts in Bangladesh: a cluster-randomised controlled trial. The Lancet. 2010;375(9721):1193–202. doi: 10.1016/s0140-6736(10)60142-0.

9. Tripathy P, Nair N, Barnett S, Mahapatra R, Borghi J, Rath S, et al. Effect of a participatory intervention with women’s groups on birth outcomes and maternal depression in Jharkhand and Orissa, India: a cluster-randomised controlled trial. The Lancet. 2010;375(9721):1182–92.

10. Fottrell E, Azad K, Kuddus A, Younes L, Shah S, Nahar T, et al. The effect of increased coverage of participatory women’s groups on neonatal mortality in Bangladesh: A cluster randomized trial. JAMA Pediatrics. 2013;167(9):816-25. Epub May 20, 2013. doi: 10.1001/jamapediatrics.2013.2534.

11. Tripathy P, Nair N, Sinha R, Rath S, Gope RK, Rath S, et al. Effect of participatory women’s groups facilitated by Accredited Social Health Activists on birth outcomes in rural eastern India: a cluster-randomised controlled trial. Lancet Glob Health. 2016;4(2):e119-28. Epub 2016/01/30. doi: 10.1016/S2214-109X(15)00287-9. PubMed PMID: 26823213.

12. Pulkki-Brannstrom AM, Haghparast-Bidgoli H, Batura N, Colbourn T, Azad K, Banda F, et al. Participatory learning and action cycles with women’s groups to prevent neonatal death in low-resource settings: A multi-country comparison of cost-effectiveness and affordability. Health Policy Plan. 2021;35(10):1280-9. Epub 2020/10/22. doi: 10.1093/heapol/czaa081. PubMed PMID: 33085753; PubMed Central PMCID: PMCPMC7886438.

13. Sinha RK, Haghparast-Bidgoli H, Tripathy PK, Nair N, Gope R, Rath S, et al. Economic evaluation of participatory learning and action with women’s groups facilitated by Accredited Social Health Activists to improve birth outcomes in rural eastern India. Cost Eff Resour Alloc. 2017;15(1):2. doi: 10.1186/s12962-017-0064-9.

14. Colbourn T, Pulkki-Brännström A-M, Nambiar B, Kim S, Bondo A, Banda L, et al. Cost-effectiveness and affordability of community mobilisation through women’s groups and quality improvement in health facilities (MaiKhanda trial) in Malawi. Cost Eff Resour Alloc. 2015;13(1).

15. UN Inter-Agency Group for Child Mortality Estimation. Levels and Trends in Child Mortality: Report 2020.. New York: United Nations Children’s Fund; World Health Organization; The World Bank; United Nations, Department of Economic and Social Affairs, Population Division; United Nations Economic Commission for Latin America and the Caribbean, Population Division, 2020.

16. Prost A, Colbourn T, Seward N, Azad K, Coomarasamy A, Copas A, et al. Women’s groups practising participatory learning and action to improve maternal and newborn health in low-resource settings: a systematic review and meta-analysis. The Lancet. 2013;381(9879):1736–46.

17. CIFF. SUSTAINABLE CHILD HEALTH SOLUTIONS: FOR THE COMMUNITY, BY THE COMMUNITY: Children Investment Fund Foundation; 2019 [cited 2021 20 Jan]. Available from: https://ciff.org/news/sustainable-child-health-solutions-community-community/.

18. EKJUT. http://www.ekjutindia.org/ 2021 x[cited 2021 01/04/2021].

19. Nair N, Tripathy PK, Gope R, Rath S, Pradhan H, Rath S, et al. Effectiveness of participatory women’s groups scaled up by the public health system to improve birth outcomes in Jharkhand, eastern India: a pragmatic cluster non-randomised controlled trial. BMJ Glob Health. 2021;6(11). Epub 2021/11/05. doi: 10.1136/bmjgh-2021-005066. PubMed PMID: 34732513; PubMed Central PMCID: PMCPMC8572384. https://www.indiacensus.net. Jharkhand Population: Estimated population in 2021 2021 [cited 2021 10 November]. Available from: https://www.indiacensus.net/states/jharkhand.

20. Ministry of Home Affairs GoI. Special Bulletin on Maternal Mortality in India 2015-2017. New Delhi: Office of the Registrar General & Census Commissioner, 2019.

21. IIPS, ICF. National Family Health Survey (NFHS-4), 2015–16: India. Mumbai: International Institute for Population Sciences 2017.

22. Scott K, George AS, Ved RR. Taking stock of 10 years of published research on the ASHA programme: examining India’s national community health worker programme from a health systems perspective. Health Res Policy Syst. 2019;17(1):29. Epub 2019/03/27. doi: 10.1186/s12961-019-0427-0. PubMed PMID: 30909926; PubMed Central PMCID: PMCPMC6434894.

23. Welfare GoIMoHaF. Home-based newborn care: operational guidelines. New Delhi2014. Available from: http://www.nhm.gov.in/index1.php?lang=1&level=3&sublinkid=1182&lid=364.

24. Murray CJL, Ezzati M, Flaxman AD, Lim S, Lozano R, Michaud C, et al. GBD 2010: design, definitions, and metrics. The Lancet. 2012;380(9859):2063–6. doi: 10.1016/S0140-6736(12)61899-6.

25. Johns B, Baltussen R, Hutubessy R. Programme costs in the economic evaluation of health interventions. Cost Eff Resour Alloc. 2003;1(1):1. PubMed PMID: 12773220; PubMed Central PMCID: PMCPMC156020.

26. Tan-Torres Edejer T BR, Adam T, Hutubessy R, Acharya A, Evans DB, Murray CJ. Making choices in health: WHO guide to cost-effectiveness analysis. Geneva: World Health Organization; 2003.

27. Wilkinson T, Sculpher MJ, Claxton K, Revill P, Briggs A, Cairns JA, et al. The International Decision Support Initiative Reference Case for Economic Evaluation: An Aid to Thought. Value Health. 2016;19(8):921-8. Epub 2016/12/19. doi: 10.1016/j.jval.2016.04.015. PubMed PMID: 27987641.

28. PPP conversion factor, GDP (LCU per international $) - India [Internet]. 2020 [cited 17/10/2020]. Available from: https://data.worldbank.org/indicator/PA.NUS.PPP?locations=IN.

29. Husereau D, Drummond M, Petrou S, Carswell C, Moher D, Greenberg D, et al. Consolidated Health Economic Evaluation Reporting Standards (CHEERS)--explanation and elaboration: a report of the ISPOR Health Economic Evaluation Publication Guidelines Good Reporting Practices Task Force. Value Health. 2013;16(2):231–50. doi: 10.1016/j.jval.2013.02.002. PubMed PMID: 23538175.

30. Lisa A. Robinson, James K. Hammitt, Michele Cecchini, Kalipso Chalkidou, Karl Claxton, Maureen Cropper, et al. Reference Case Guidelines for Benefit-Cost Analysis in Global Health and Development. 2019.

31. WHO. Global Health Observatory data repository: Life Tables WHO; 2019 [cited 2021 01/08/2021]. Available from: https://apps.who.int/gho/data/view.main.60740?lang=en.

32. Haacker M, Hallett TB, Atun R. On discount rates for economic evaluations in global health. Health Policy Plan. 2020;35(1):107-14. Epub 2019/10/19. doi: 10.1093/heapol/czz127. PubMed PMID: 31625564.

33. Woods B, Revill P, Sculpher M, Claxton K. Country-Level Cost-Effectiveness Thresholds: Initial Estimates and the Need for Further Research. Value in Health. 2016;19(8):929–35. doi: 10.1016/j.jval.2016.02.017. PubMed PMID: PMC5193154.

34. Tan-Torres Edejer T, Baltussen R, Adam T, Hutubessy R, Acharya A, Evans DB, et al., editors. Making choices in health: WHO guide to cost-effectiveness analysis. Geneva: World Health Organization; 2003.

35. Gupta S, Kumar N, Menon P, Pandey S, and Raghunathan K. Engaging women’s groups to improve nutrition: Findings from an evaluation of the JEEViKA multisectoral convergence pilot in Saharsa, Bihar. Washington, DC: World Bank., 2019.

36. Chandrashekar S, Saha S, Varghese B, Mohan L, Shetty G, Porwal A, et al. Cost and cost-effectiveness of health behavior change interventions implemented with self-help groups in Bihar, India. PLoS One. 2019;14(3):e0213723. Epub 2019/03/29. doi: 10.1371/journal.pone.0213723. PubMed PMID: 30921334; PubMed Central PMCID: PMCPMC6438566.

37. Siwach G, de Hoop T, Ferrari G, Yulia. B. Guidelines on Estimating the Costs and Cost-Effectiveness of Women’s Groups in International Development. The Evidence Consortium on Women’s Groups (ECWG), 2019.

38. Maitra C, Hodge A, Jimenez Soto E. A scoping review of cost benefit analysis in reproductive, maternal, newborn and child health: What we know and what are the gaps? Health Policy Plan. 2016;31(10):1530-47. Epub 2016/11/02. doi: 10.1093/heapol/czw078. PubMed PMID: 27371550.

